# Economic Evaluation of State Control, Low Price, and Research-Based Policy for Eating Disorders Treatment in Sweden

**DOI:** 10.1101/2023.05.30.23290679

**Authors:** Per Södersten, Ulf Brodin, Cecilia Bergh

**Author notes:** Corresponding author: Per Södersten, Karolinska Institutet, the Mandometer Clinic, Novum, S-141 04 Huddinge, Sweden.

## Abstract

**Introduction:** Porter and Olmsted Teisberg suggested that the value of healthcare should be measured by treatment outcomes related to costs. Policy should financially reward treatment effects and in an outline of value-based healthcare, they predicted:

1. State control policy yields variable effects, increasing costs
2. Low price policy decreases effects, increasing costs
3. Research and Development (R&D) policy increases effects, decreasing costs

**Methods:** The treatment of eating disorders in Region Stockholm, Sweden, in years 2012-2016 makes it possible to test these predictions because a State control, a Low price, and an R&D provider were contracted and the effects and costs of their treatments are publically available from Region Stockholm.

**Results:** The State control provider was contracted to provide more care services than the other providers. The average yearly number of patients treated to remission/patients treated was 164/714 (23%) at the State control provider, 41/170 (24) at the Low price provider, and 152/192 (79%) at the R&D provider. The average yearly budget was 73 million Swedish crowns (MSEK) compared to 49 MSEK at the R&D provider and 32 MSEK at the Low price provider (on average 30% lower per care service than the two other providers). The average cost to treat a patient to remission/patients treated was highest at the Low price provider (859 KSEK), followed by the State control provider (464 KSEK) and the R&D provider (327 KSEK).

**Conclusions:** The results confirm Porter and Olmsted Teisberg’s three predictions and suggest that an R&D policy increases the value of healthcare.

## Introduction

Porter and Olmsted Teisberg suggested that the value of healthcare should be measured by treatment outcomes related to cost,^1^ and outlined three policy-based, cost-effect relationships in value-based healthcare:

1. State control policy yields variable effects, increasing costs
2. Lo price policy decreases effects, increasing costs
3. Research and Development (R&D) policy increases effects, decreasing costs

Sweden provides unique opportunities for evaluating treatment outcomes because of its National Quality Registries, but value-based healthcare met resistance,^2^ and was not considered by policy makers. However, the three relationships can be tested in retrospect because Region Stockholm contracted three providers of eating disorders treatment, that were governed by each of these policies in years 2012-2016.

## Methods

The providers were contracted to perform six main, individually priced care services (eTable). The associated effect (i.e. number of patients treated to remission) and cost at the State control provider,^3^ the Low price provider,^4^ and the Research and Development (R&D) provider^5^ for years 2012-2016 are publically available from Region Stockholm. The economic value of the policies was calculated as the cost for treating a patient to remission relative to all patients treated.

## Results

The contracted prices at the State control provider were similar to those at the R&D provider and about 30% lower at the Low price provider (eTable).

Because it was contracted to perform more care services than the other providers, the State control provider treated almost four times as many patients (Figure, panel 1), but a variable number of patients to remission, and on average only 11 patients (7%) more than the R&D provider (panel 2). The Low price provider treated fewer patients to remission, on average 27% of those treated to remission by the R&D provider (panel 2). The relationship between patients treated to remission and patients treated was on average 23% at the State control provider, 24% at the Low price provider, and 79% at the R&D provider.

**Figure:**
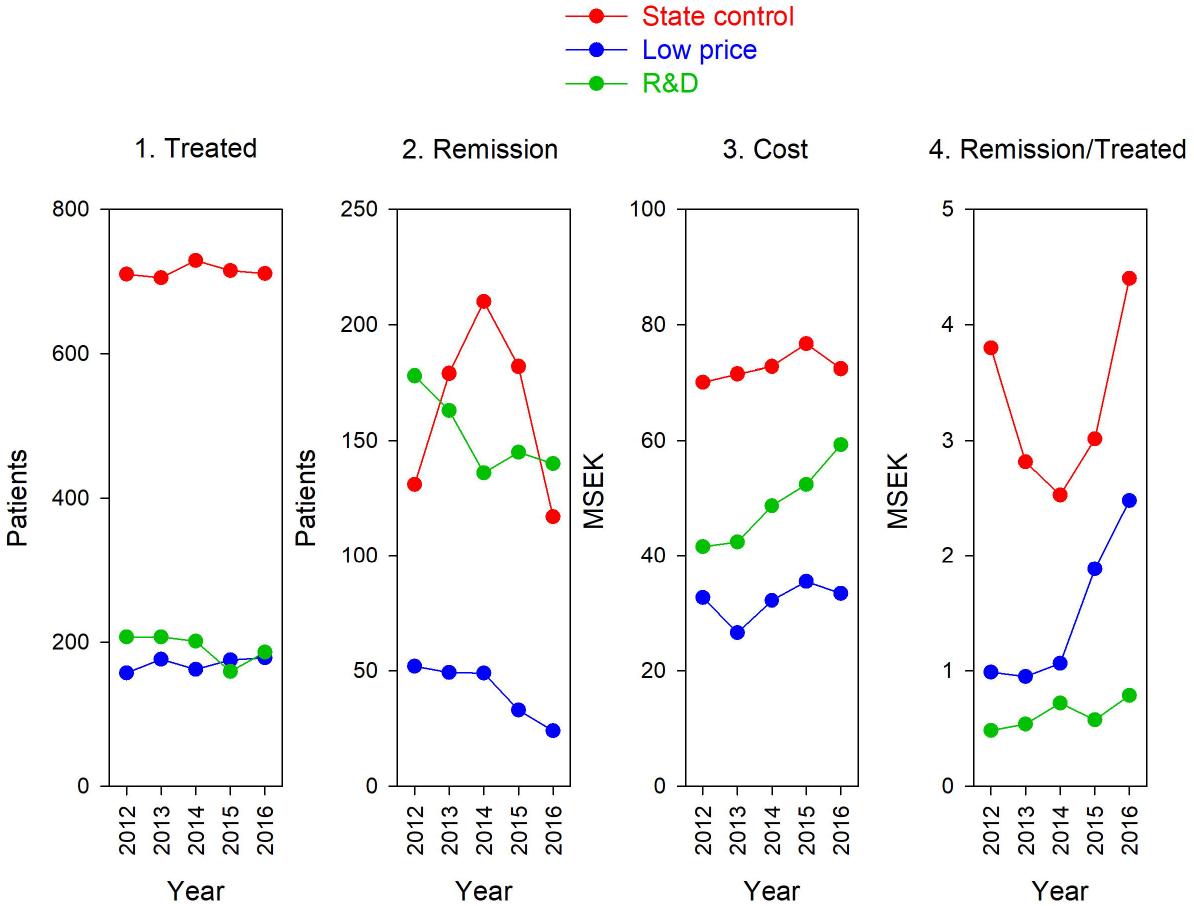
Effect and cost of State control, Low price, and R&D policy on the treatment of patients with eating disorders in Region Stockholm. Number of patients treated (panel 1), number of patients treated to remission (panel 2), cost for treating all patients (panel 3, million Swedish crowns, MSEK), and cost for treating a patient to remission in relation to all patients treated (panel 4).

The annual cost at the State control provider was the highest and relatively stable over the years (panel 3). The cost was lowest at the Low price provider, and higher and increasing at the R&D provider (panel 3).

Because of the differences in outcomes, the cost to treat a patient to remission at the State control provider was variable and high, increased over time at the Low price provider, and increased somewhat but was lowest at the R&D provider (panel 4).

## Discussion

Here we confirmed the prediction that a policy based on R&D yields a higher value of healthcare than policies based on state control or low price, by treating more patients to remission. Economic evaluation based on outcome unveiled the “paradox” that low price is expensive,^1^ and that state control offers little control because only a few patients are treated to remission; quite a few must have dropped out of treatment relatively quickly.

Today, effect measures are used in less than a fifth of public procurements of healthcare services in Sweden, the emphasis is on a low price, obscuring the value of healthcare services.^6^ The results presented here support the original suggestion that the value of healthcare “can only be measured at the treatment level”.^1^ Policies with no mechanism for reward (state control) of based on fee-for-service (low price) provide less value.

A limitation of this study is that it was made possible as the result of a political decision, not by design. Designing such a study would, however, be difficult ethically. A strength is that the patients were admitted to all clinic by the same criteria and treated over the same period of time.

## Data Availability

All data produced in the present work are contained in the manuscript

## Acknowledgement

Mando Group AB supported this work financially. The economic analysis was presented at the Association for Psychological Science: International Convention of Psychological Science (ICPS), Brussels, March 9-11, 2023. Poster number III-108.

