## Supplementary information for "Economic Evaluation of State Control, Low Price, and Research-Based Policy for Eating Disorders Treatment in Sweden"

**eTable.** Contracted Prices for Six Care Services at the State Control, the Low Price, and the R&D Provider of Eating Disorders Treatment in Region Stockholm

**Contracted Prices for Eating Disorders Treatment**

Region Stockholm contracted three providers of eating disorders treatment, based on three policies: State control, Low price, and Research and Development (R&D).

The main effect of the treatment, remission (absence of an eating disorders diagnosis), has been reported to Region Stockholm^1^ and information is available for years 2012-2016.^2^

Contracted prices for years 2012-2016 are available from Region Stockholm.^1^

All providers were paid for six main care services (eTable). Region Stockholm also contracted the providers separately for other care services and adjusted payments on a yearly basis, comparison between these differences in costs is not possible.

**eTable. Contracted Prices for Six Care Services at the State Control, the Low Price, and the R&D Provider of Eating Disorders Treatment in Region Stockholm**

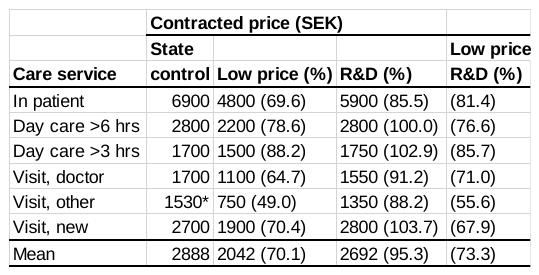

Abbreviation: SEK, Swedish Crowns

*Average of three visits: by relative (1900 SEK), by a group (800 SEK, and in a network (1900 SEK)
